# Tract-based white matter hyperintensity patterns in patients with Systemic Lupus Erythematosus using an unsupervised machine learning approach

**DOI:** 10.1101/2022.03.04.22271909

**Authors:** Theodor Rumetshofer, Francesca Inglese, Jeroen de Bresser, Peter Mannfolk, Olof Strandberg, Andreas Jönsen, Anders Bengtsson, Markus Nilsson, Linda Knutsson, Jimmy Lätt, Gerda M. Steup-Beekman, Tom W.J. Huizinga, Mark A. van Buchem, Itamar Ronen, Pia C. Sundgren

## Abstract

Currently, little is known about the spatial distribution of white matter hyperintensities (WMH) in the brain of patients with Systemic Lupus erythematosus (SLE). Previous lesion markers, such as number and volume, ignore the strategic location of WMH. The goal of this work was to develop a fully-automated method to identify predominant patterns of WMH across WM tracts based on cluster analysis. A total of 221 SLE patients with and without neuropsychiatric symptoms from two different sites were included in this study. WMH segmentations and lesion locations were acquired automatically. Cluster analysis was performed on the WMH distribution in 20 WM tracts. Our pipeline identified five distinct clusters with predominant involvement of the forceps major, forceps minor, as well as right and left anterior thalamic radiations and the right inferior fronto-occipital fasciculus. The patterns of the affected WM tracts were consistent over the SLE subtypes and sites. Our approach revealed distinct and robust tract-based WMH patterns within SLE patients. This method could provide a basis, to link the location of WMH with clinical symptoms. Furthermore, it could be used for other diseases characterized by presence of WMH to investigate both the clinical relevance of WMH and underlying pathomechanism in the brain.

## 1. INTRODUCTION

White matter hyperintensities (WMH) are lesions in the white matter (WM) appearing hyperintense on T2-weighted MRI images^1^. The prevalence of WMH in the general population increases with age^1^. WMH are considered as important neuroimaging and clinical markers in many neurological diseases^2^. However, the pathogenesis of WMH is not well understood and may have various etiologies. The prevalence of WMH is highly variable within and across different diseases^2,3^. WMH are one of the main imaging findings observed in the brain in systemic lupus erythematosus (SLE) patients^4^, even though not all SLE patients manifest WMH^5,6^. SLE is a rare autoimmune disease affecting mostly women and it is characterized by the production and deposition of several autoantibodies. SLE involves different organs, including the central nervous system, in which damage could lead to neuropsychiatric (NP) syndromes^6^. The American Colleague of Rheumatology (ACR) describes 19 NP syndromes that could be present in SLE patients. These ACR criteria also subdivide SLE patients experiencing NP events in two subgroups based on the attribution of NP events directly related to the disease (NPLSE) or other causes (non-NPSLE)^7,8^. In clinical practice, however, the attribution process of the NP events to the disease is difficult as the nature of NP syndromes is highly heterogeneous and ranges from mild (e.g. headache and anxiety) to major symptoms (e.g. seizures and psychosis)^9^. In addition, there are large differences in the NP attribution across sites and across studies (between 37% and 95%)^4^, illustrating the difficulty in establishing a rigorous standard for the attribution of NP to SLE^10^. We used the term NPSLE to refer to patients with NP manifestations attributed to SLE and the term non-NPSLE to patients with NP manifestations not attributed to SLE.

The origin of WMH in SLE is not fully understood, but could be the result of inflammatory and immunologically mediated small vessel disease (SVD)^3^. The location of WMH in white matter has been shown to be of major importance in several neurological diseases. In multiple sclerosis (MS) and Alzheimer’s disease it has been demonstrated that WMH location is more strongly linked to neuropsychological impairment than WMH volume and count^11,12,13^. WMH location in patients with arterial diseases can provide prognostic survival information^14^. WMH in SLE patients has not been shown to be homogeneous across subgroups^15^. One study reported that the prevalence of WMH in the splenium of the corpus callosum, in the right superior longitudinal fasciculus and in some small clusters in the right corona radiata was higher in NPSLE patients compared to SLE patients without NP involvement^16^. Another study reported high WMH burden in the superior longitudinal fasciculus and anterior corona radiata in NPSLE as well as in SLE patients without NP involvement^17^. All previous assessments of WMH burden on specific white matter tracts in SLE were based on manual WMH segmentation. Although manual segmentation is often considered as gold standard, it inevitably introduces variability that can be reduced by devising an automated WMH segmentation pipeline^18,19^. The variability in results obtained from WMH segmentation algorithms compared to manual segmentation, seems to depend on WMH burden. However, existing WMH segmentation algorithms are robust for a wide range of WMH load^20,21,22^ and a fully automated pipeline could increase the level of reproducibility.

The goal of our retrospective cross-sectional study was to develop a fully automated method to characterize the spatial distribution of WMH across WM tracts in SLE patients. Due to the well-known heterogeneity in diagnosis and difficulties in the attribution of NP manifestations, we aimed for a highly objective approach by investigating only the spatial distribution of WMH in the brain. We used an unsupervised machine learning method to identify clusters based on WM tract-based abnormalities. We addressed the typical paucity of subjects in SLE studies by pooling two cohorts of SLE patients. Further, we investigated if our method is robust across SLE subgroups and clinical and radiological differences between the sites.

## 2. METHODS

### 2.1 Subject population

#### 2.1.1 Leiden cohort

The Leiden University Medical Center (LUMC) is the national referral center in the Netherlands for SLE patients experiencing NP symptoms. The SLE patients undergo a one-day standardized evaluation that includes multidisciplinary medical assessments and complementary tests, including extensive laboratory tests, neuropsychological testing and a brain MRI scan^23,24^. All patients are assessed by a rheumatologist, neurologist, psychiatrist, vascular internal medicine expert and advanced nurse practitioner. This evaluation is followed by a multidisciplinary consensus meeting in order to decide whether the NP events are attributable to SLE or not. In the final attribution of NP symptoms to SLE or other etiologies, several aspects are taken into account: the time between the onset of NP symptoms and diagnosis of SLE, SLE disease activity, the type of NP symptoms, favoring factors and the presence of alternative diagnoses^10,25^. NPSLE diagnoses were defined according to the 1999 ACR nomenclature^26^. All patients fulfilled the 1997 revised ACR criteria for the classification of SLE^27^.

From this cohort, 216 patients scanned between May 2007 and April 2015 were eligible. Information on sex, age, disease duration and age of disease onset was obtained via interview with the patient and retrieved from electronical medical records. SLE activity and damage indices were scored for each patient: the SLE disease activity was determined using the Systemic Lupus Erythematosus Disease Activity Index 2000 (SLEDAI-2K)^28^; SLE irreversible damage was assessed with the Systemic Lupus International Collaborating Clinics/American College of Rheumatology damage index (SDI)^8^. The Leiden-The Hague-Delft ethics approval committee approved the study (registration number P07.177) and all included patients signed informed consent.

All participants were scanned using a Philips Achieva 3T MRI scanner (Philips Healthcare, Best, The Netherlands) equipped with a body transmit RF coil and an 8-Channel receive head coil array. A standardized scanning protocol was used. The sequences included in this study were: a 3D T1-weighted scan (voxel size = 1.17 × 1.17 × 1.2 mm^3^; TR/TE = 9.8/4.6 ms) and two versions of a fluid-attenuated inversion recovery (FLAIR) scan. A total of 99 data sets included a 2D-multislice FLAIR sequence (voxel size = 1.0 × 1.0 × 3.6 mm^3^; TR/TE/TI = 10000/120/2800 ms) and 53 data sets included a 3D FLAIR (voxel size = 1.10 × 1.11 × 0.56 mm^3^; TR/TE/TI = 4800/576/1650 ms) (see Supplementary Table S1 for a summary of the MRI methods). The change from 2D to 3D in the FLAIR protocol occurred in February 2013.

#### 2.1.2 Lund cohort

SLE patients experiencing NP symptoms were recruited by the Department of Rheumatology in Lund, Skåne University Hospital, Sweden. Inclusion criteria were: female sex, age between 18 and 55 years and right handedness. Patients with any contraindication to MRI or pregnancy were not asked to participate in this study. All patients fulfilled the SLICC classification criteria for SLE^29^. Extensive laboratory and neuropsychological testing were performed in the Lund cohort as well. The brain MRI scan was performed on the same day as the clinical visit with few exceptions due to logistical issues (maximum of 2 weeks in difference, e.g. patients requested another time slot). The collected clinical data and the NP symptoms, as defined by the American College of Rheumatology (ACR) case definition for NPSLE^26^, were evaluated by a rheumatologist and a neurologist. In case of split opinions, a consensus meeting followed.

In the Lund cohort, 73 subjects, recruited consecutively from January 2013 to January 2016, were eligible for this study. All participants underwent rheumatologic and standardized neurologic clinical assessment. Information about SLE disease activity and organ damage were recorded according to the SLE disease Activity Index 2000 (SLEDAI-2k)^28^ and the Systemic Lupus Erythematosus International Collaborating Clinics / ACR Damage Index (SLICC/ACR-DI)^8^. The Regional Ethical Review Board in Lund, Sweden (#2012/4, #2014/748) approved this study and a written informed consent was obtained for all subjects prior to inclusion.

All participants underwent a brain scan on a 3T MRI Siemens scanner (Siemens MAGNETON Skyra, Erlangen, Germany). Imaging protocols included in this study were: T1-weighted magnetization-prepared rapid gradient-echo (MPRAGE) (1 mm isotropic voxels, TR/TE = 1900/2.54 ms) and 2D-multislice T2-weighted FLAIR (0.7×0.7×3.0 mm, TR/TE/TI = 9000/81/2500 ms) (see Supplementary Table S1).

### 2.2 Cluster analysis

The preprocessing of image data, prior to cluster analysis is shown in Figure 1. A detailed description of all preprocessing steps can be found in the Supplementary Material (Material Section). For the cluster analysis, the WMH burden on each WM tract was L2-normalized (unit norm) to obtain an individual WMH pattern for each subject. All subjects were included in this analysis without giving any information about clinical diagnose, site or NP manifestations. Hierarchical clustering (Ward’s method)^30^ was applied to the L2-normalized WMH load from 20 WM tracts (based on the JHU WM atlas) and a total of 186 SLE patients, resulting in 186 feature vectors with 20 values. SLE patients without detectable WMH (n=35) were not included in the clustering but were included in the statistics. The cluster analysis and the performance evaluation of the clustering procedure was performed with scikit-learn 0.20.3 (RRID:SCR_002577)^31^.

**Figure 1.**
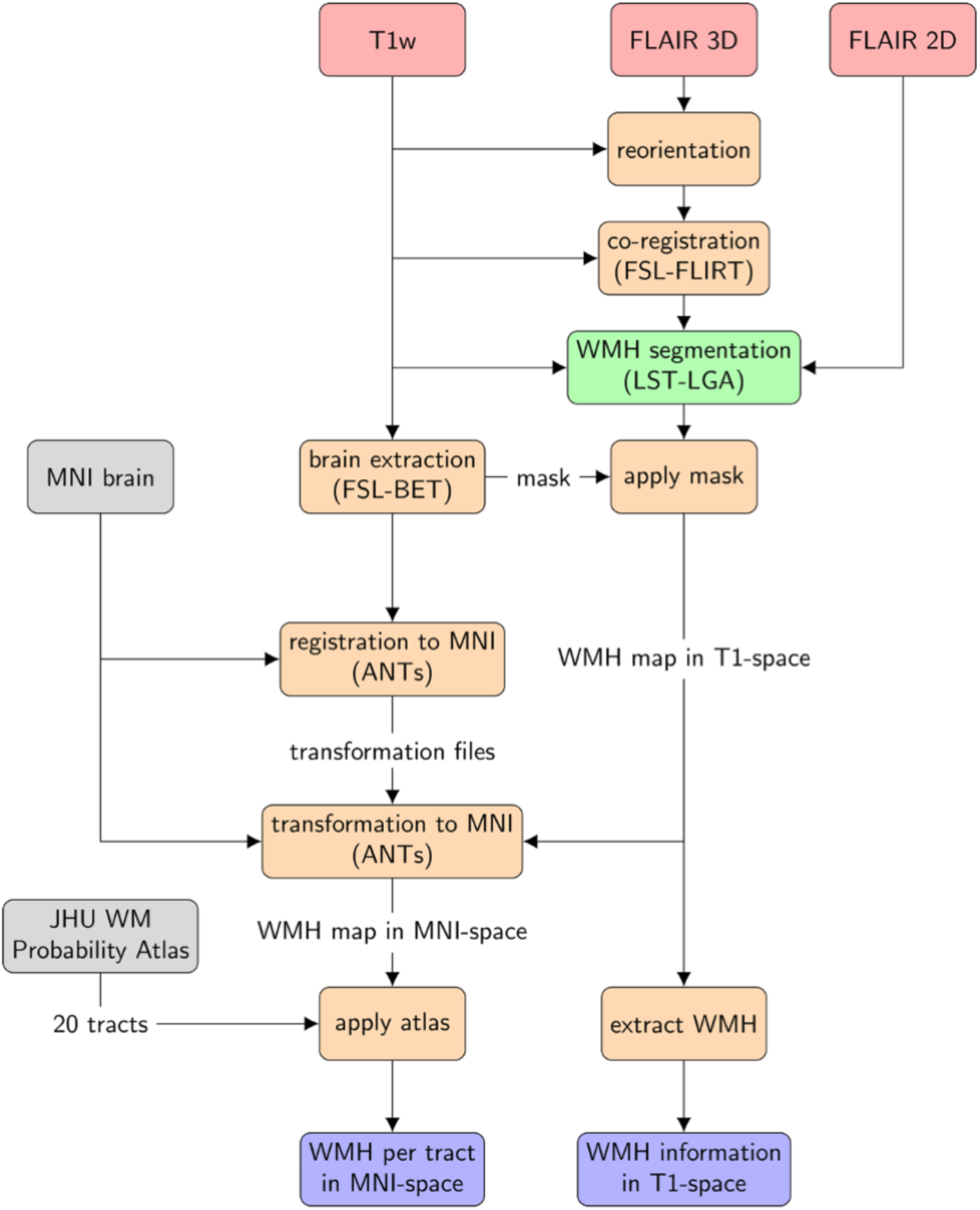
Preprocessing workflow. Workflow of the fully automated approach. 3D-FLAIR images are reoriented and co-registered to the T1-weighted (T1w) before WMH segmentation. White matter hyperintensities (WMH) segmentation is performed with the Lesion Segmentation Toolbox-Lesion Growth Algorithm (LST-LGA) using T1w and FLAIR images. The volume and number of WMH are extracted from the WMH maps in T1-space. The WMH probability maps are transformed to Montreal Neurological Institute (MNI) space by applying the transformation from the T1w images. Those maps are masked by the Johns Hopkins University (JHU) white matter (WM) probability atlas to obtain the tract specific WMH volumes. To quantitatively assign WMH to specific WM tracts the probability values of superimposed voxels on the lesion map and the WM tract are multiplied and the resulting product is summed over the entire tract. FLAIR= fluid-attenuated inversion recovery; FSL= FMRIB Software Library; FLIRT= Linear Image Registration Tool; BET= Brain Extraction Tool; ANTs= Advanced Normalization Tools

Agglomerative hierarchical clustering successively merges groups of subjects (starting with each subject in its own group) based on the Euclidean distance between their WMH feature vector, until all subjects form a single cluster. The successive merging of subgroups results in a tree structure, or *dendrogram*, shown in Supplementary Figure S1. The iterative calculation of inter-cluster distances was computed using Ward’s method, resulting in minimal intra-cluster variance. Each node or branching point of the tree corresponds to the merging of two clusters, for which the corresponding inter-cluster distance is shown on the y-axis. To estimate the optimal number of clusters, the dendrogram has to be cut at a certain distance threshold. The optimal cluster number was evaluated by a consensus of two different methods: Silhouette Coefficient^32^ and the Calinski-Harabasz index^33^.

To evaluate the robustness of the method, three sensitivity analyses were performed. In the first one, cluster analysis was performed on each SLE subgroup. The second sensitivity analysis was implemented separately on the Lund and Leiden cohorts. The last sensitivity analysis was performed by clustering the total SLE patients using as a regressor the site (Lund, Leiden) including: sex, type of FLAIR (2D, 3D), age, disease duration, SDI-score, SLEDAI-2k-score, and WMH total volume.

### 2.3 Statistical analysis

The between-group and cohort differences in demographic and clinical data, were assessed for nominal variables with Chi-square test and for continuous variables based on their non-normal distribution with Mann-Whitney U test. Differences between clusters were estimated for nominal variables with Chi-square test and for continuous variables with non-parametric Kruskal-Wallis test. Post-hoc pairwise comparisons of the clusters were performed with Mann-Whitney U-test and using Bonferroni multiple comparison correction. Data are represented as number (percentage) or median (10-90 percentile). All statistical analyses as well as tests for distribution normality (D’Agostino and Pearson’s test) were performed using Python package Scipy 1.2.1 (RRID:SCR_008058)^34^. Covariates correction was performed with a general linear model (GLM) using Python package statsmodels 0.10.1 (RRID:SCR_016074)^35^.

## 3. RESULTS

### 3.1 Demographic and clinical data

From the Leiden cohort, 216 patients were eligible for this study. Of these, 28 patients were excluded because of undefined and mixed subgroups, 8 for misdiagnosis established during a follow-up visit, 3 patients for motion artefacts in the MRI scans, 3 patients for co-registration failure of the T1 and FLAIR images using the LST-LGA toolbox, 20 patients for the presence of brain infarcts over 1.5 cm that hindered accurate brain volume measurements and 2 patients were removed due to the presence of other diseases (brain tumor and large arachnoid cyst). This resulted in a total of 152 patients included in the present study, comprising 37 NPSLE and 115 non-NPSLE patients.

From the Lund cohort, 73 subjects were eligible for this study. A total of 4 subjects were excluded due to misdiagnosis, temporal lobe resection, hypothyroidism and co-registration failure of the LST-LGA toolbox. In total, 69 patients were included in the present study, comprising 42 NPSLE and 27 non-NPSLE.

Table 1 shows clinical and demographic data of the two different cohorts (Leiden/NL and Lund/SWE). Statistically significant differences were observed in sex, age, disease duration, age of onset, SLE disease scores, ACR criteria, volume and number of WMH (p<0.05).

**Table 1.**
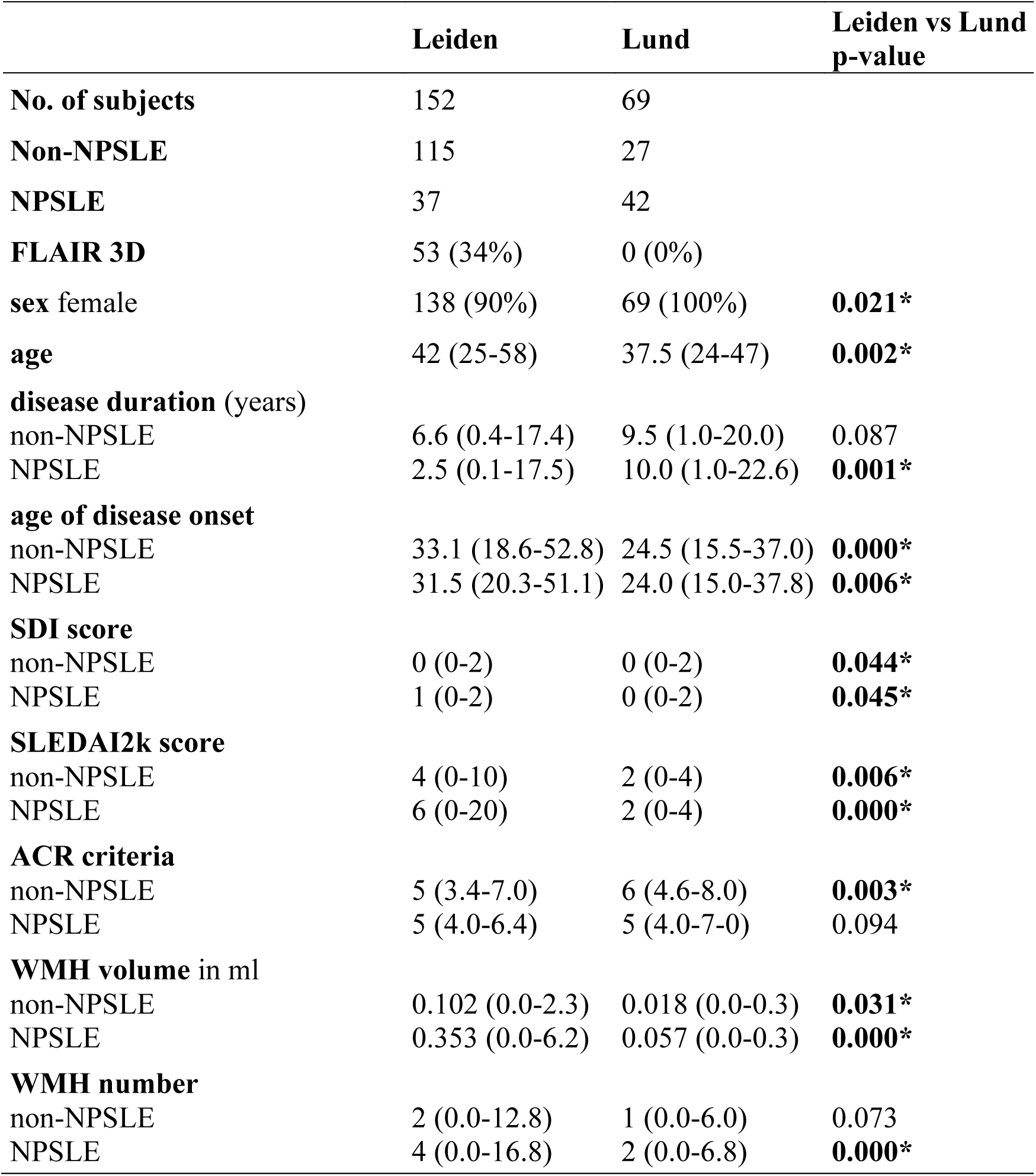
Demographic and clinical data across the two site cohorts. Data are represented as number (percentage) or median (10-90 percentile). Differences between Leiden and Lund cohort are expressed in p-value and calculated for nominal variables with Chi-square tests (sex) and for continuous variables, based on their not-normally distribution, with Mann-Whitney U tests. NPSLE= neuropsychiatric systemic lupus erythematosus; SDI= systemic lupus international collaborating clinics damage index; SLEDAI-2K= systemic lupus erythematosus disease activity index 2000; ACR= American College of Rheumatology. *= p value<0.05

|  | Leiden | Lund | Leiden vs Lund<br>p-value |
| --- | --- | --- | --- |
| **No. of subjects** | 152 | 69 |  |
| **Non-NPSLE** | 115 | 27 |  |
| **NPSLE** | 37 | 42 |  |
| **FLAIR 3D** | 53 (34%) | 0 (0%) |  |
| **sex female** | 138 (90%) | 69 (100%) | **0.021\*** |
| **age** | 42 (25-58) | 37.5 (24-47) | **0.002\*** |
| **disease duration (years)** |  |  |  |
| non-NPSLE | 6.6 (0.4-17.4) | 9.5 (1.0-20.0) | 0.087 |
| NPSLE | 2.5 (0.1-17.5) | 10.0 (1.0-22.6) | **0.001\*** |
| **age of disease onset** |  |  |  |
| non-NPSLE | 33.1 (18.6-52.8) | 24.5 (15.5-37.0) | **0.000\*** |
| NPSLE | 31.5 (20.3-51.1) | 24.0 (15.0-37.8) | **0.006\*** |
| **SDI score** |  |  |  |
| non-NPSLE | 0 (0-2) | 0 (0-2) | **0.044\*** |
| NPSLE | 1 (0-2) | 0 (0-2) | **0.045\*** |
| **SLEDAI2k score** |  |  |  |
| non-NPSLE | 4 (0-10) | 2 (0-4) | **0.006\*** |
| NPSLE | 6 (0-20) | 2 (0-4) | **0.000\*** |
| **ACR criteria** |  |  |  |
| non-NPSLE | 5 (3.4-7.0) | 6 (4.6-8.0) | **0.003\*** |
| NPSLE | 5 (4.0-6.4) | 5 (4.0-7.0) | 0.094 |
| **WMH volume in ml** |  |  |  |
| non-NPSLE | 0.102 (0.0-2.3) | 0.018 (0.0-0.3) | **0.031\*** |
| NPSLE | 0.353 (0.0-6.2) | 0.057 (0.0-0.3) | **0.000\*** |
| **WMH number** |  |  |  |
| non-NPSLE | 2 (0.0-12.8) | 1 (0.0-6.0) | 0.073 |
| NPSLE | 4 (0.0-16.8) | 2 (0.0-6.8) | **0.000\*** |

An overview of the cerebrovascular risk factors, ongoing pharmacologic treatments as well as antibodies can be found in Supplementary Table S2.

From both cohorts, a total of 221 data sets were analyzed in this study, of which 79 were NPSLE and 142 were non-NPSLE. Supplementary Table S3 includes demographic and clinical data for the two subgroups. No significant differences were found between the two groups in demographic and clinical variables.

### 3.2 Cluster analysis

The cluster analysis was performed on all patients and yielded in five distinct clusters (Figure 2): cluster 1 (n=52) is mainly assigned to the forceps major and to lesser extent to the left and right inferior fronto-occipital fasciculus; cluster 2 (n=57) mainly to the right anterior thalamic radiation and to lesser extent to the forceps minor and the right inferior fronto-occipital fasciculus; cluster 3 (n=23) to the forceps minor; cluster 4 (n=30) to the left anterior thalamic radiation and to lesser extent to the forceps minor and the right anterior thalamic radiation; andcluster 5 (n=24) was more heterogeneous in terms of location in WM tracts but it could be mainly assigned to the right inferior fronto-occipital fasciculus. A total of 35 patients (8 NPSLE, 27 non-NPSLE) with no WMH were excluded from cluster analysis. The lesion frequency maps (Figure 3) show the WMH location probability in each cluster.

**Figure 2.**
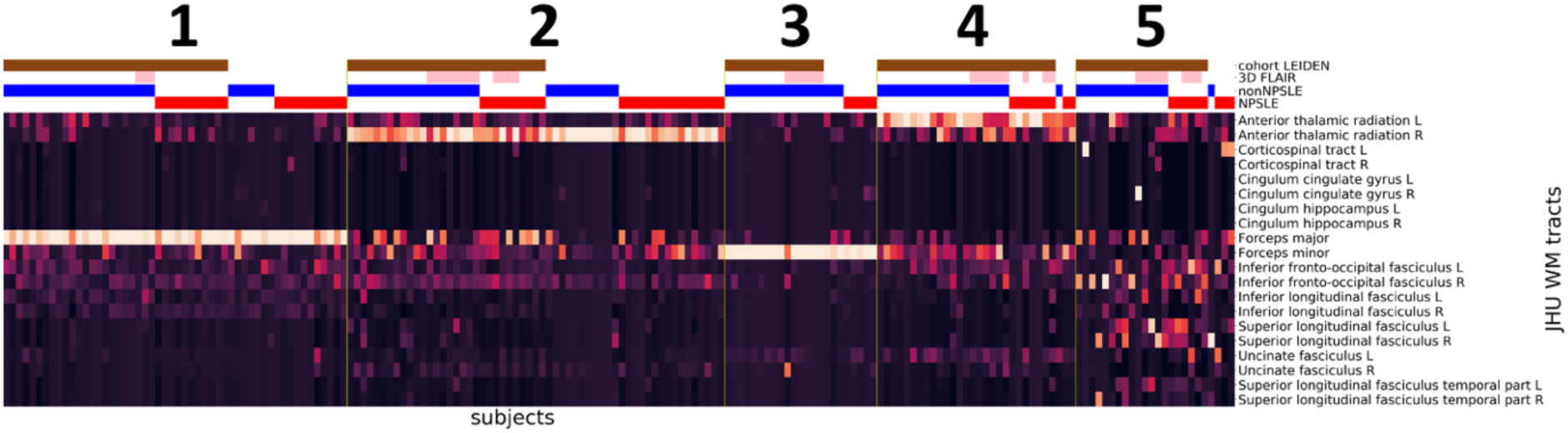
Cluster analysis on the entire SLE cohort. Heatmaps showing the 5 different MRI subtypes after the hierarchical clustering with the L2-normalization was performed. Subjects are shown on the x-axis and the Johns Hopkins University (JHU) white matter probability atlas tracts on the y-axis. The horizontal bars at the top show additional information: Leiden cohort (brown) complemented by the Lund cohort, 3D-FLAIR (pink) complemented by 2D-FLAIR, non-NPSLE (blue), NPSLE (red). FLAIR= fluid-attenuated inversion recovery; NPSLE= neuropsychiatric systemic lupus erythematosus.

**Figure 3.**
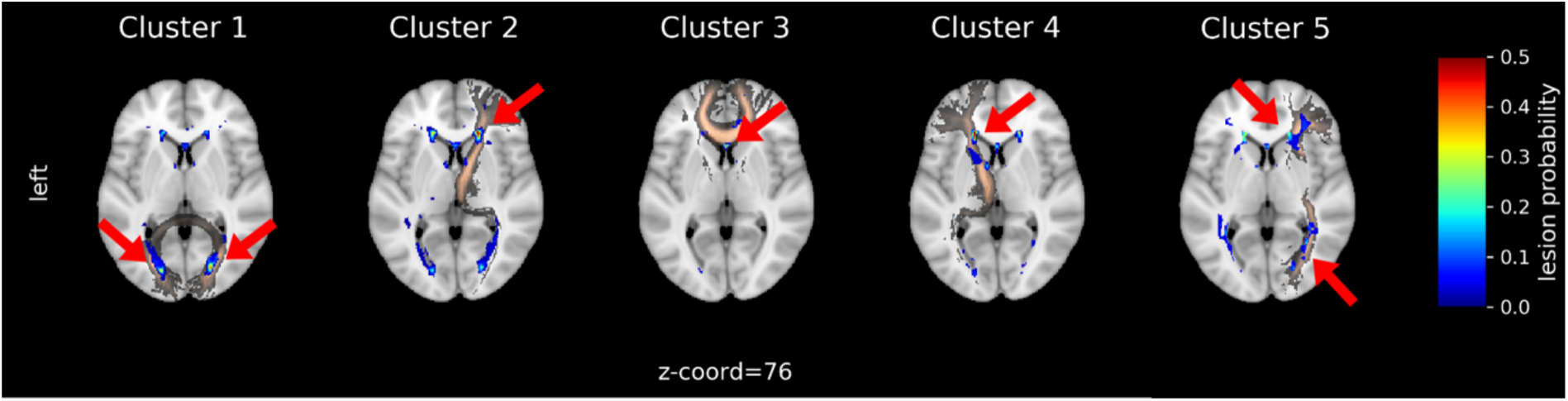
Lesion frequency map for each cluster in MNI-space. WMH in cluster 1 can be mainly assigned to Forceps Major, cluster 2 to right Anterior Thalamic Radiation, cluster 3 to Forceps Minor and 4 to the left Anterior Thalamic Radiation. Cluster 5 shows a high WMH burden and can be assigned to the right inferior fronto-occipital fasciculus. Clusters are shown as lesion probabilities from 0.0 to 0.5 (color scale on the right). The main WMH corresponding to specific WM tracts (copper color) are emphasized with red arrows.

Patient age, volume and number of WMH lesions were statistically significantly different across clusters (p=0.005, p=0.008 and p=0.003 respectively). After correction for multiple testing, patient age was significantly higher (p=0.002) in cluster 5 compared to patients without detectable WMH. WMH volume and number in cluster 4 was significantly higher than in cluster 3 (p<0.001) and number of lesions was significantly higher in cluster 2 compared to cluster 3 (p=0.001) (Supplementary Table S4).

### 3.3 Performance evaluation of the cluster analysis

The cluster analysis was evaluated through a consensus of two different methods. The Silhouette coefficient and Calinski-Harabaz index showed a clear peak using five clusters (Figure 4). Although the Silhouette coefficient seems to increase with higher cluster size (with local maximum for 5 clusters), the Calinski-Harabaz index sharply decreases with increasing number of clusters, therefore n(clusters) = 5 was chosen for best performance.

**Figure 4.**
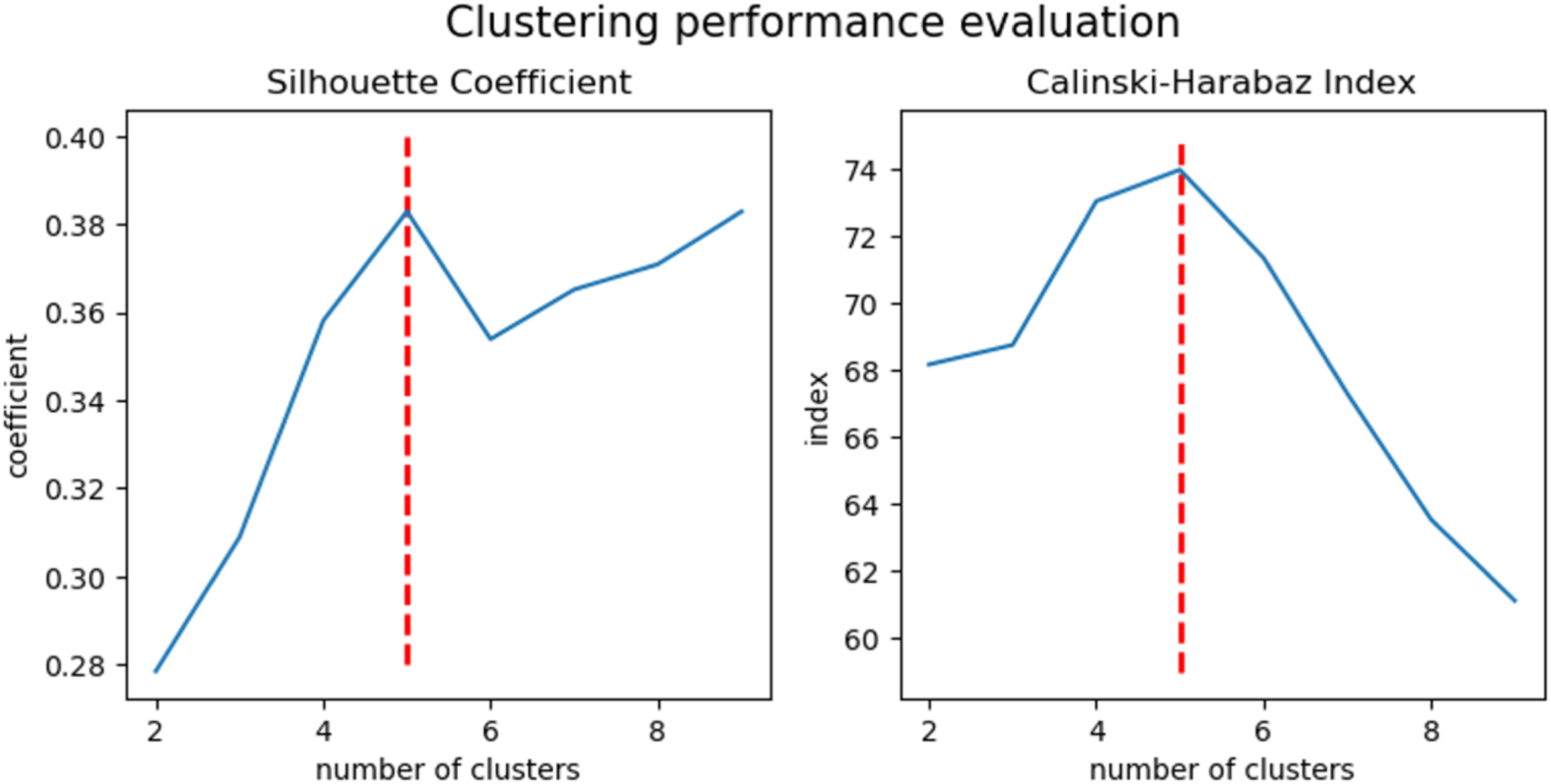
Clustering performance evaluation. To determine the optimal number of clusters in the hierarchical cluster analysis a consensus of two different methods were used. The mean Silhouette Coefficient (on the left) is calculated over mean intra-cluster distance divided by the minimal inter-cluster distance to the nearest cluster. Positive values indicate a dense clustering whereas negative an incorrect clustering. The Calinski-Harabaz index (on the right) is the ratio of the sum of distances squared between and within the clusters. A high index indicated a dense and well separated cluster. Both methods indicate an optimal number of clusters of 5.

### 3.4 Sensitivity analysis

Sensitivity analysis was implemented in three different ways. The first sensitivity analysis was performed separately on each of the two subgroups of SLE patients: NPSLE and non-NPSLE. This resulted in an overlap of the number of the same patients included in a certain cluster of 83% for the NPSLE group and 85% for the non-NPSLE group, when compared to the main analysis where all SLE patients were included. The cluster analysis on NPSLE patients resulted in 6 clusters. Similarly to the cluster analysis performed on the entire group, clusters 1, 2, 3 and 5 were mainly assigned to specific WM tracts: forceps major, right anterior thalamic radiation, forceps minor and left anterior thalamic radiation, respectively. Cluster 4 consisted of two subjects and was attributed to the left inferior fronto-occipital fasciculus. Cluster 6 was more heterogeneous in terms of WM tract location and was comparable to cluster 5 of the main analysis (Figure 5). The corresponding NP manifestations in the NPSLE subgroup showed no correlation with the unveiled clusters (Supplementary Figure S2). The cluster analysis on non-NPSLE patients revealed 5 clusters. Clusters 1, 2, 3 and 4 could be mainly assigned to the same specific WM tracts identified in the main cluster analysis: forceps major, forceps minor, right anterior thalamic radiation and left anterior thalamic radiation, respectively. Similar to the NPSLE subgroup clustering, cluster 5 was more heterogeneous in terms of WM tract location and comparable to cluster 5 of the main analysis (Figure 5).

**Figure 5.**
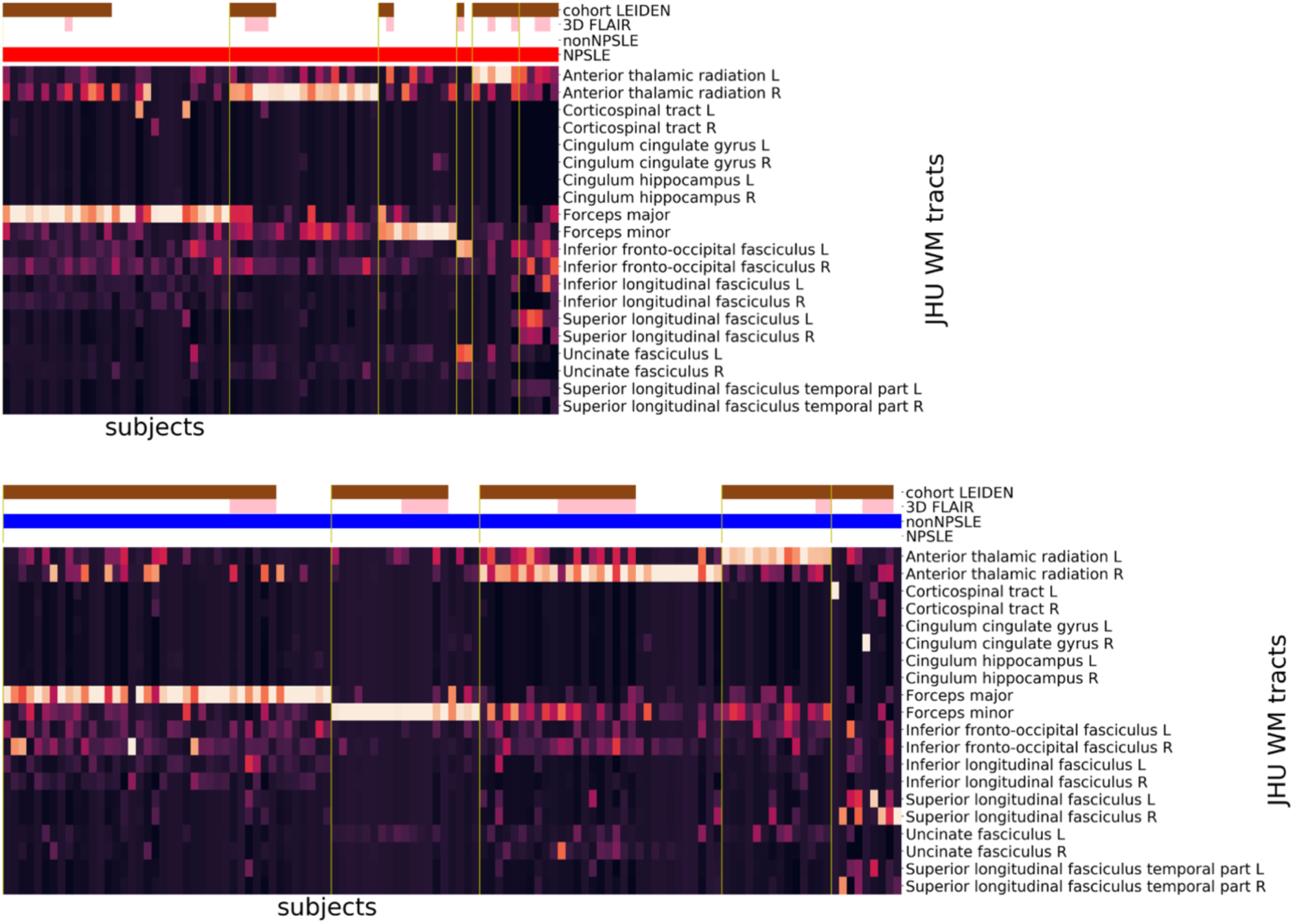
Sensitivity analysis on NPSLE and non-NPSLE patients. Cluster analysis on NPSLE patients (top) and on non-NPSLE patients (bottom). The horizontal bars at the top show additional information: Leiden cohort (brown) complemented by the Lund cohort, 3D-FLAIR (pink) complemented by 2D-FLAIR, non-NPSLE (blue), NPSLE (red). FLAIR= fluid-attenuated inversion recovery; NPSLE= neuropsychiatric systemic lupus erythematosus.

The second sensitivity analysis was performed on each site separately (Figure 6). Compared to the main analysis, the overlap of the same patients in the certain clusters resulted in 93% for the Lund cohort and 87% for the Leiden cohort. The four clusters revealed from clustering the Lund cohort separately, can be assigned to the right anterior thalamic radiation, the forceps minor, the forceps major, similar to the main analysis, and a heterogenous cluster in terms of the affected tracts, similar to the cluster 5 in the main analysis. The five clusters revealed from clustering the Leiden cohort separately can be assigned to the forceps major, the forceps minor and the left and right anterior thalamic radiation, similar to the main analysis, and a heterogenous cluster in terms of the affected tracts, similar to cluster 5 in the main analysis.

**Figure 6.**
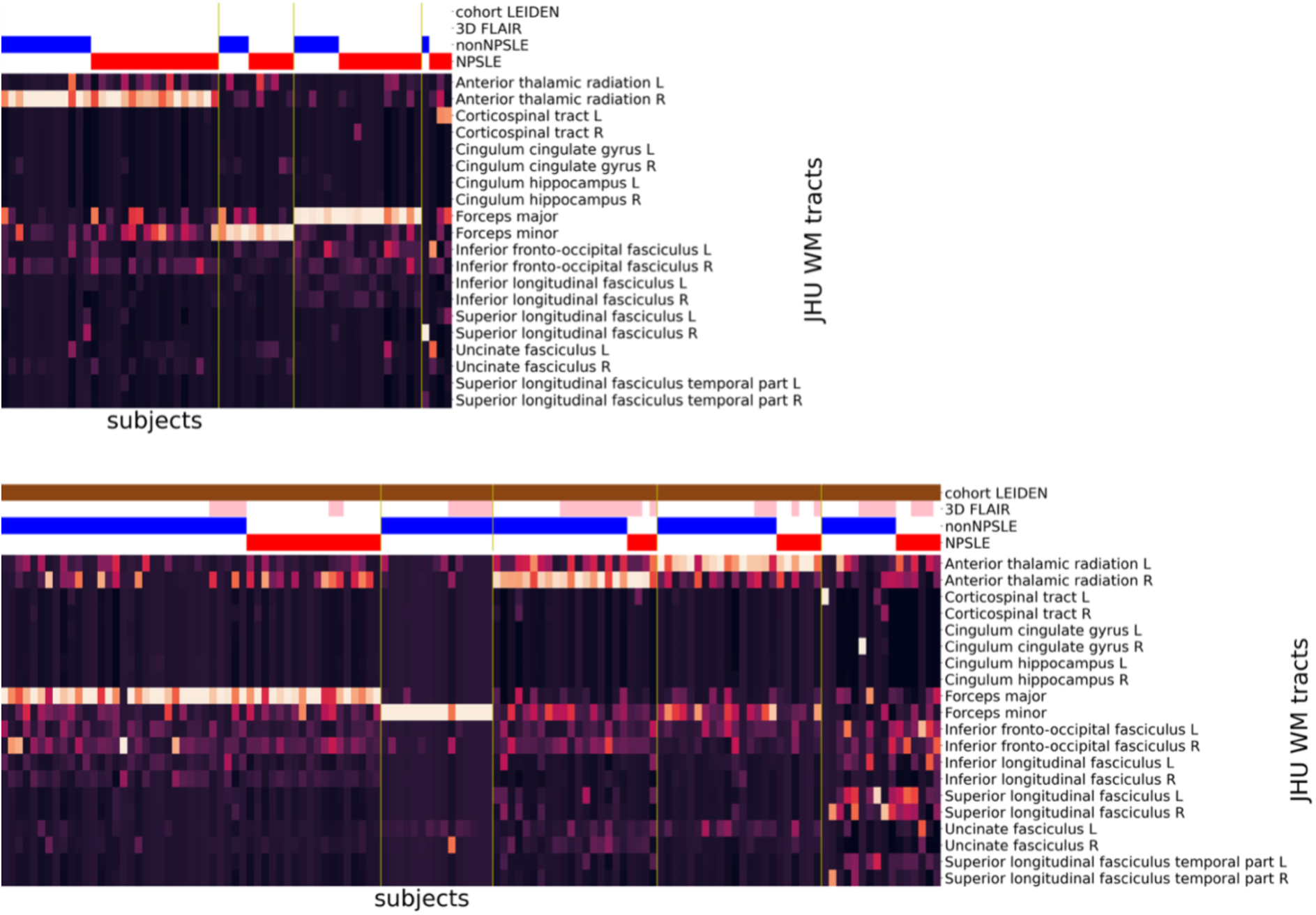
Sensitivity analysis on each cohort separately. Cluster analysis on Lund (top) and Leiden cohort (bottom). The horizontal bars at the top show additional information: Leiden cohort (brown) complemented by the Lund cohort, 3D-FLAIR (pink) complemented by 2D-FLAIR, non-NPSLE (blue), NPSLE (red). FLAIR= fluid-attenuated inversion recovery; NPSLE= neuropsychiatric systemic lupus erythematosus

The third and last sensitivity analysis was performed on the entire SLE population using as regressors the site and the significant differences between the cohorts: sex, type of FLAIR, age, disease duration, SDI-score, SLEDAI-2k-score and the WMH total volume using a general linear model (GLM). The resulting clusters showed an overlap of 90% with those obtained by the main analysis (Figure 7). Cluster 1 to 4 could be mainly assigned to specific WM tracts: forceps major, right anterior thalamic radiation, forceps minor and left anterior thalamic radiation, respectively. Cluster 5 was more heterogeneous in terms of WM tract location and was comparable to cluster 5 of the main analysis.

**Figure 7.**
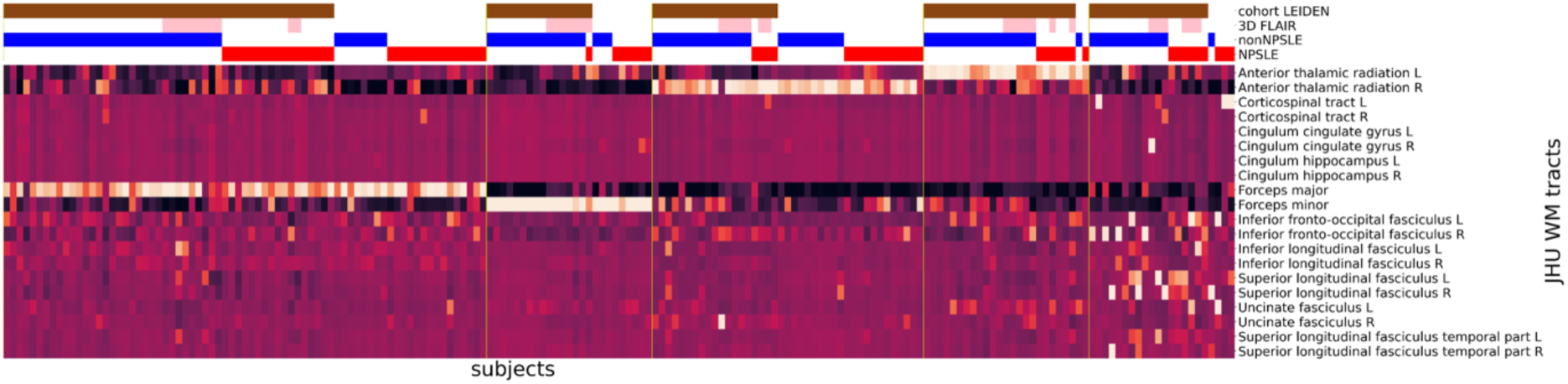
Sensitivity Analysis using GLM. Cluster analysis on the entire SLE cohort performed after L2 normalization and GLM model to correct for cohort, type of FLAIR, sex, age, disease duration. SDI-score, SLEDAI-2k-score and WMH total volume. The horizontal bars at the top show additional information: Leiden cohort (brown) complemented by the Lund cohort, 3D-FLAIR (pink) complemented by 2D-FLAIR, non-NPSLE (blue), NPSLE (red). GLM= General linear model; FLAIR= fluid-attenuated inversion recovery; NPSLE= neuropsychiatric systemic lupus erythematosus; SDI= systemic lupus international collaborating clinics damage index; SLEDAI-2K = systemic lupus erythematosus disease activity index 2000.

### 3.5 Volumetric analysis of the WMH

The distributions of the WMH volumes on each WM tract for each cluster on the total SLE population are given in Supplementary Table S5. With the exception of the right corticospinal tract and the left cingulum cingulate gyrus, statistically significant differences in WMH volumes were found in all WM tracts, across all five clusters. The main WM tract assigned to each cluster shows also the highest volume.

The WMH volume was significantly higher in NPSLE patients compared to non-NPSLE patients in the right anterior thalamic radiation (p=0.024), in the right inferior fronto-occipital fasciculus (p=0.010), in the right inferior longitudinal fasciculus (p=0.041) and in the right uncinate fasciculus (p=0.033) (Supplementary Table S6).

## 4. DISCUSSION

We developed a fully automated method to unveil WMH pattern in SLE patients experiencing NP events. We applied this method on a two-center dataset of SLE patients. Our method detected five robust and distinct clusters, characterized by the involvement of the forceps major, forceps minor as well as the left and right anterior thalamic radiation and the right inferior fronto-occipital fasciculus (Figures 2 and 3). Our results are consistent across the two subgroups, NPSLE and non-NPSLE patients (Figure 4). Despite the heterogeneity of the disease, our results are consistent across both sites (Figure 6) and are not affected by the clinical and radiological differences between the two cohorts (Figure 7). Differences in volume and number of WMHs were observed between the clusters and subgroups as presented in the Supplementary Tables S5 and S6.

Cluster analysis has been applied successfully to identify distinct clusters based on coarse location of WMH in other brain disorders, such as arterial disease^36^ and postoperative delirium^37^. So far, unsupervised machine learning approaches based on structural MRI information were not explored in research involving SLE patients^5,6^. To the best of our knowledge, the work we present here is the first machine learning analysis that focuses on brain features gauged by MRI in SLE. Compared to previous studies^16,17^, in our developed method the WMH were detected and assigned to WM tracts automatically by using a well-established lesion segmentation algorithm and further processed by publicly available software^38^. Further, the L2-normalization highlighted the underlying WMH pattern for each patient by reducing the impact of the total WMH burden and harmonizing the two-sites dataset. These steps, in combination with a machine learning technique unveiled a consistent spatial pattern of the mainly affected WM tracts. Few studies applied cluster analysis in SLE but those are based on clinical features, such as demographic^39^, genetic^40^ and autoantibodies^41^ data.. In contrast, our developed approach focuses on MRI brain features which could provide a basis to link neuroimaging findings to clinical symptoms.

Several studies in other diseases, such as Alzheimer’s disease and MS showed the importance to categorize spatially WMH to the link with neuropsychological impairment^12,13^. Previous studies in SLE patients showed higher prevalence of WMH on specific WM tracts^16,17,42,43,44^. However, manual segmentation of the WMH and their subsequent ways to assign WMH to specific WM tracts may have influenced the reproducibility of the study and make it difficult to compare them with our approach. Furthermore, these studies included also SLE patients without any NP syndromes, a subgroup which was not included in the present study^16,17^. Our method identified a set of WM tracts with the highest lesion volume, which seem to be those most significantly involved in our SLE patients experiencing NP manifestations. In healthy elderly, WMH on tracts adjacent to the frontal horns of lateral ventricles, such as the left and right anterior thalamic radiation, are associated with worse performance in executive function^45,46^ and planning complex behavior^46^. Indeed, decrease of complex planning behavior performance is shown in SLE patients^47^. Microstructural WM abnormalities in forceps minor are higher in patient with Schizophrenia compared to controls^48^ and are related to depression and fatigue in MS^49^. Furthermore, both anterior thalamic radiation and forceps minor are linked to cognitive impairment in patients with SVD^50^. Since the importance of the frontal WM tracts in cognition and psychiatric disorder, future studies are needed to fully understand the role that the tracts we found in this study may play in the performance of NPSLE patients.

All SLE patients, except those without detectable WMH, were included in the cluster analysis without considering the subgroups or the affiliation to cohorts. The attribution process of the NP events is difficult since the nature of NP syndromes is heterogeneous (can vary from headache and seizures to anxiety and psychosis)^9^ and there are large differences in the NP attribution across studies (between 37% and 95%)^4^. It has been demonstrated that misclassification of NP events may occur in clinical practice with an over attribution of NP to the disease (NPSLE)^51^. Several challenges are related to the diagnostic procedure of NPSLE. First, SLE is categorized as a rare disease (prevalence 1-5/10 000, source www.orpha.net) and many SLE studies suffer from small sample sizes^5,52^, hampers to draw conclusions regarding the reliability and robustness of results. To overcome this problem, we performed a two site study. Second, the absence of biomarkers (radiological or laboratory) reliable enough in the diagnostic process make it difficult to create a link between radiological findings and clinical symptoms (clinic-radiological paradox)^5^.

In this study we could not find an association between the NP manifestations and the clusters in the NPSLE subgroup. We assume that the reason for this lies in the strong variability of NP manifestations between the two clinical cohorts (Supplementary Figure S2) and in the highly heterogenous nature of NP syndromes. Even though NPSLE patients showed higher WMH volume in some WM tracts compared to non-NPSLE patients, the cluster analysis showed that the WM tracts most affected by WMH are similar in both subgroups. This suggests that location and pattern of WMH have no correlation with NPSLE diagnosis and attribution to the disease. Therefore, location may give a new important information about WMH in SLE. Describing WMH only in terms of volume or number may not give enough information about etiology but in severity. Furthermore, despite the heterogeneity of the NP events within each site and the diagnostic, clinical and radiological variability across sites, our results appeared to be robust and stable. This is the first comprehensive study to examine WMH in SLE that assesses a broad categorization of WMH in terms of load, location and volume using a fully automated method in two site cohort.

Our work is not without limitations. Some clusters identified by our method comprise a low number of patients. This is expected, since SLE is a rare disease, and even with our effort to increase the patient population by merging data from two centers, the overall number of patients remains small compared to similar studies performed in more common diseases. A significant limitation of this study is the lack of a subgroup of SLE patients without NP symptoms. The lack of such a group stems from the retrospective nature of this study and the lack of availability of imaging data within our database. Recruiting such a cohort in future studies will undoubtedly strengthen any conclusion regarding the link between spatial distribution of brain abnormalities and NP symptoms in SLE, as it has been repeatedly shown that patients with SLE without NP also exhibit brain abnormalities, albeit to a lesser extent^53^. Further, the focus on structural MRI information omits the possible impact of clinical features, such as the presence of clinical activity, antiphospholipid antibodies positivity as well as cardiovascular risk factors and other factors that were not included in the present analysis. This is a limitation of this study, and future multicenter studies would benefit from incorporating such data in the analysis. Additionally, patients for which automated detection of WMH yielded no positive results, were not included in our analysis. This study was retrospectively performed in prospective cohorts, therefore differences in MRI protocol, diagnosis definition, and clinical data are present and can contribute to biases between groups and to an increased variance. The MRI sequences, and in particular the FLAIR sequences, were different between sites. However, in all our analyses, subgroups, cohorts and 3D FLAIR scans were not predominant in a specific cluster. Differences in NP attribution and diagnosis between the two sites, could have had an effect on the results. However, the sensitivity analysis we performed show that the WMH patterns obtained by our clustering strategy are robust even when SLE subgroups and cohorts are clustered separately, or when corrected for significant clinical and radiological differences.

To conclude, we developed a method based on an unsupervised machine learning approach and identified a WMH pattern which was consistent in a two-site cohort. With our approach, we provided a fully automated standardized method to identify tract-based WMH patterns. The identification of affected WM tracts via the clustering algorithm was robust, despite heterogeneity of the NP events and their association with the disease. Allocation of the WMH burden to the most affected WM tracts could help investigate the link between radiological findings and clinical symptoms in SLE patients with NP manifestations. In a future study an association between pathogenesis, overall phenotypes and even genetics would be interesting to explore.

## Data Availability

Under General Data Protection Regulation (GDPR) restrictions, the MRI and other patient data cannot be made publicly available. Each of the two sites involved in this study (Lund University, Leiden University Medical Center) can share anonymized data with individual sites following an appropriate data transfer agreement (DTA). Under such agreement, each site (Lund, Leiden) will share only their own data with the signing site. Requests for DTA should be sent to Pia Sundgren (Lund University) or Itamar Ronen (Leiden University Medical Center).
The code used for data preprocessing and cluster analysis can be found in the following GitHub repository: https://github.com/TheoRum/manuscript_WMH_SLE

https://github.com/TheoRum/manuscript_WMH_SLE

## AUTHOR CONTRIBUTIONS

**TR:** Software, Data curation, Formal analysis, Visualization, Roles/Writing – original draft. **FI:** Data curation, Visualization, Formal analysis, Roles/Writing – original draft. **JdB:** Roles/Writing – original draft, Validation. **PM:** Data curation, Software, Writing – review and editing. **AJ:** Resources, Validation, Writing – review and editing. **AB:** Resources, Validation, Writing – review and editing. **MN:** Conceptualization, Validation, Writing – review and editing. **LK:** Conceptualization, Validation, Writing – review and editing. **JL:** Data curation, Software, Writing – review and editing. **GMSB:** Resources, Validation, Writing – review and editing. **TWJH:** Resources, Validation, Writing – review and editing. **MAvB:** Validation, Writing – review and editing, **OS:** Conceptualization, Software, Writing – review and editing. **IR:** Funding acquisition, Supervision, Project administration, Roles/Writing – original draft. **PCS:** Funding acquisition, Supervision, Project administration, Roles/Writing – original draft.

## DATA AVAILABILITY STATEMENT

Under General Data Protection Regulation (GDPR) restrictions, the MRI and other patient data cannot be made publicly available. Each of the two sites involved in this study (Lund University, Leiden University Medical Center) can share anonymized data with individual sites following an appropriate data transfer agreement (DTA). Under such agreement, each site (Lund, Leiden) will share only their own data with the signing site. Requests for DTA should be sent to Pia Sundgren (Lund University) or Itamar Ronen (Leiden University Medical Center).

The code used for data preprocessing and cluster analysis can be found in the following GitHub repository: https://github.com/TheoRum/manuscript_WMH_SLE

## ACKNOWLEDGEMENTS

The study was supported by funding by Regional Research founds (RegSkane 625631), SUS Foundation and Donation funds (PCS), Alfred Österlund foundation (PCS), Swedish Rheumatism Association R-56371 (PCS), King Gustaf V’s 80-year foundation (FAI-2017-0341 and FAI-2019-0559) (PCS).

The funding sources had no involvement in the study design, in the collection, analysis and interpretation of data, in the writing of the report and in the decision to submit the article for publication.

## ETHICS STATEMENT

All patients included in this study signed informed consent and it was approved by the Leiden-The Hague-Delft, The Netherlands, ethics approval committee (registration number P07.177) and the Regional Ethical Review Board in Lund, Sweden (#2012/4, #2014/748).

