## Supplementary for "Tract-based white matter hyperintensity patterns in patients with Systemic Lupus Erythematosus using an unsupervised machine learning approach"

**Preprocessing of MRI data**

Preprocessing of image data, prior the cluster analysis, was performed using Nipype 1.5.1 (RRID:SCR_002502). The preprocessing pipeline is shown in Figure 1. $T_1$-weighted images were initially brain-extracted using the Brain Extraction Tool (BET) in FMRIB Software Library (FSL) software 5.0.10 (RRID:SCR_002823)[1] and registered to MNI space (1mm isometric) using Advanced Normalization Tools (ANTs) 1.9.2 (RRID:SCR_004757)[2].
Subsequently, all 3D-FLAIR scans were reoriented, resampled and co-registered to the 3D-$T_1$-weighted images using the Linear Image Registration Tool (FLIRT) of (FSL 5.0.10)[3,4]. Those steps were only necessary for the 3D-FLAIR scans, due to co-registration errors in the fully automated WMH segmentation toolbox. Segmentation was performed using the Lesion Segmentation Toolbox-Lesion Growth Algorithm (LST-LGA) 3.0 included in the statistical parametric mapping (SPM12) software (RRID:SCR_007037)[5]. LST-LGA resampled and co-registered the FLAIR images to the $T_1$-weighted images before WMH segmentation, i.e. all FLAIR images resulted in the same resolution. The generated WMH probability maps were binarized using the default initial threshold (kappa) of 0.3 to extract the number and volume of the WMH. This was done using an in-house developed Python script. To reduce the impact of false positives and small non-specific lesions the WMH maps were thresholded to probability greater than 0.5, and lesions smaller than 0.015 ml were removed. The original WMH probability maps were transformed to MNI-space by applying the same transformation matrices as those obtained from registration of the $T_1$-weighted images. No additional smoothing was applied since the transformations already included such processing steps.
As a last step, to quantitatively assign WMH volumes to specific WM tracts, lesion maps were masked by the Johns Hopkins University (JHU) WM tract probability atlas[6], which consists of 20 WM tracts. The probability values of superimposed voxels on the WMH map and the WM tract were multiplied and the resulting product was summed over the entire tract. Further multiplying the probabilities by voxel size yielded our measure of WMH volume in each WM tract[7]. Patients without WMH, and with WMH below the above-mentioned thresholds (of lesion probability, size and without overlap to the WM tracts) were defined as patients with non-detectable WMH. In addition, a quality assessment was performed by visually evaluating all preprocessing step for each subject by trained researchers (TR and FI), and when deemed necessary by two experienced neuroradiologists (PCS and JB).

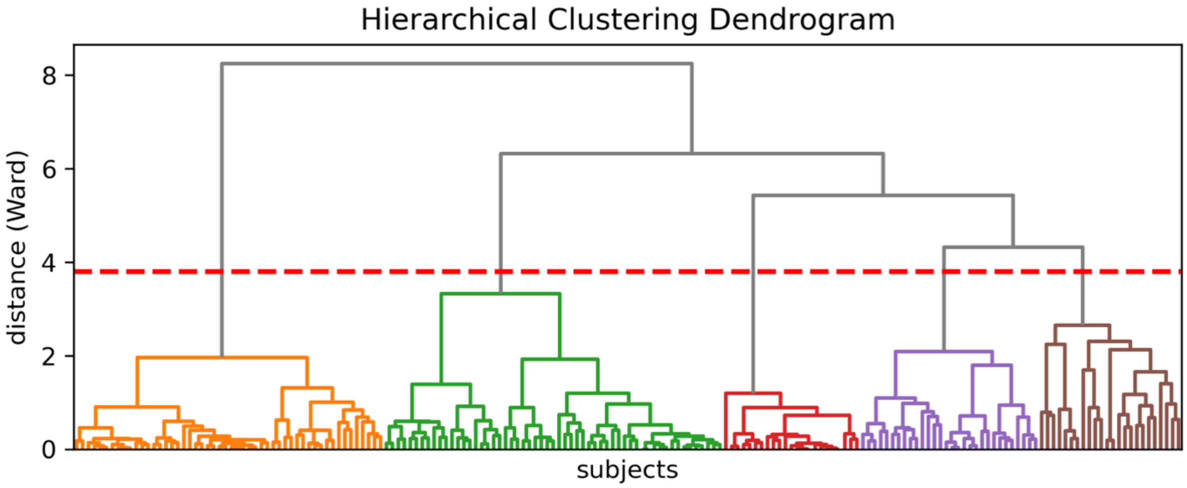


**Supplementary Figure S1. Dendrogram.** The dendrogram visualizing the cluster hierarchy result of the cluster analysis. The y-axis represents the Euclidean distance between the subjects using Wards' method and subjects or groups of subjects are linked together with horizontal lines. The x-axis shows the different subjects colored in the resulted clusters. The red dashed line indicates the optimal cut-off point for five clusters.

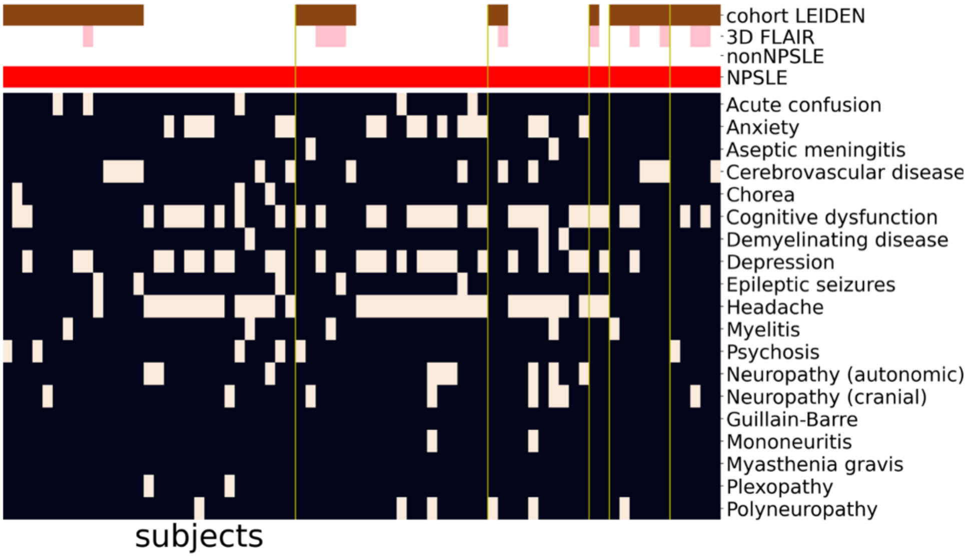


**Supplementary Figure S2. NP manifestations NPSLE clustering.** Corresponding neuropsychiatric syndromes of the separate clustering of the NPSLE patients. The horizontal bars at the top show additional information: Leiden cohort (brown) complemented by the Lund cohort, 3D-FLAIR (pink) complemented by 2D-FLAIR, non-NPSLE (blue), NPSLE (red).

FLAIR= fluid-attenuated inversion recovery; NPSLE= neuropsychiatric systemic lupus erythematosus.

**Supplementary Table S1. MRI acquisition parameters.** MRI acquisition characteristics from both cohorts. From the Leiden cohort, FLAIR images from 99 subjects were acquired with a 2D-multislice and 53 subjects with a 3D sequence.
TR= repetition time; TE= echo time; TI= inversion time; TFE= turbo field echo; MPRAGE= magnetization prepared rapid gradient echo; FLAIR= fluid-attenuated inversion recovery.

| | **Leiden** | **Lund** |
|---|---|---|
| **MRI scanner** | 3T Philips Achieva | 3T Siemens Skyra |
| **Transmit coil** | body coil | body coil |
| **Receive coil** | 8-channel head coil | 20-channel head coil |
| **$T_1$-weighted-sequence** | 3D-TFE | MPRAGE |
| resolution | 1.17 x 1.17 x 1.2 mm | 1.0 x 1.0 x 1.0 mm |
| TR | 9.8 ms | 1900 ms |
| TE | 4.6 ms | 2.54 ms |
| **$T_2$-weighted FLAIR** | 2D | 2D |
| **sequence** | 1.0 x 1.0 x 3.6 mm | 0.7 x 0.7 x 3 mm |
| resolution | 10000 ms | 9000 ms |
| TR | 120 ms | 81 ms |
| TE | 2800 ms | 2500 ms |
| TI | | |
| | 3D | |
| | 1.10 x 1.11 x 0.56 | |
| resolution | mm | |
| TR | 4800 ms | |
| TE | 576 ms | |
| TI | 1650 ms | |

**Supplementary Table S2. Cerebrovascular risk factors, antibodies and pharmacological treatment.** Data are represented for each variable are presented as number (percentage). NPSLE= neuropsychiatric systemic lupus erythematosus; SDI= systemic lupus international collaborating clinics damage index; SLEDAI-2K = systemic lupus erythematosus disease activity index 2000; ACR= American College of Rheumatology; DMARD= disease-modifying antirheumatic drug; MMF= mycophenolate mofetil; IVIG= intravenous immunoglobulins

| | **Leiden** | **Lund** |
|---|---|---|
| **No. of subjects** | 152 | 69 |
| **Non-NPSLE** | 115 | 27 |
| **NPSLE** | 37 | 42 |
| **Smoking** | | |
| non-NPSLE | 56 (48%) | 2 (7%) |
| NPSLE | 15 (40%) | 5 (11%) |
| **Diabetes** | | |
| non-NPSLE | 6 (5%) | 0 (0%) |
| NPSLE | 2 (5%) | 0 (0%) |
| **Hypertension** | | |
| non-NPSLE | 38 (33%) | 4 (15%) |
| NPSLE | 15 (40%) | 10 (24%) |
| **Anti-Nuclear Antibodies** | | |
| non-NPSLE | 79 (68%) | 27 (100%) |
| NPSLE | 32 (86%) | 41 (97%) |
| **Anti-ds-DNA Antibodies** | | |
| non-NPSLE | 25 (21%) | 14 (51%) |
| NPSLE | 19 (51%) | 27 (64%) |
| **Anti-SM-Nuclear Antigen** | | |
| non-NPSLE | 6 (5%) | 5 (18%) |
| NPSLE | 2 (5%) | 4 (9%) |
| **Glucocorticoids** | | |
| non-NPSLE | 56 (49%) | 19 (70%) |
| NPSLE | 26 (70%) | 35 (83%) |
| **Antimalarials** | | |
| non-NPSLE | 76 (66%) | 23 (85%) |
| NPSLE | 23 (62%) | 31 (74%) |
| **Anti-Hypertensive treatment** | | |
| Non-NPSLE | 33 (29%) | 7 (26%) |
| NPSLE | 14 (38%) | 15 (36%) |
| **Cyclophosphamide** | | |
| Non-NPSLE | 2 (2%) | 0 (0%) |
| NPSLE | 2 (5%) | 0 (0%) |
| **MMF** | | |
| Non-NPSLE | 14 (12%) | 5 (19%) |
| NPSLE | 1 (3%) | 11 (26%) |

| | | |
|---|---|---|
| **Azathioprine** | | |
| Non-NPSLE | 21 (18%) | 10 (37%) |
| NPSLE | 8 (22%) | 12 (29%) |
| **Rituximab** | | |
| Non-NPSLE | 0 (0%) | 0 (0%) |
| NPSLE | 0 (0%) | 1 (2%) |
| **IVIG** | | |
| Non-NPSLE | 0 (0%) | 1 (4%) |
| NPSLE | 0 (0%) | 1 (2%) |
| **Methotrexate** | | |
| Non-NPSLE | 10 (9%) | 1 (4%) |
| NPSLE | 2 (5%) | 0 (0%) |
| **Thalidomide** | | |
| Non-NPSLE | 0 (0%) | 0 (0%) |
| NPSLE | 0 (0%) | 0 (0%) |
| **Leukeran** | | |
| Non-NPSLE | 0 (0%) | 0 (0%) |
| NPSLE | 0 (0%) | 0 (0%) |
| **Belimumab** | | |
| Non-NPSLE | 0 (0%) | 6 (22%) |
| NPSLE | 0 (0%) | 2 (5%) |
| **DMARD total** | | |
| Non-NPSLE | 47 (41%) | 23 (85%) |
| NPSLE | 13 (35%) | 27 (64%) |

**Supplementary Table S3. Demographic and clinical data for each SLE subgroup.** Data are represented as number (percentage) or median (10-90 percentile). Differences between NPSLE and non-NPSLE are expressed in p-value and calculated for nominal variables with Chi-square tests (sex) and for continuous variables, based on their not-normally distribution, with Mann-Whitney U tests.
NPSLE= neuropsychiatric systemic lupus erythematosus; SDI= systemic lupus international collaborating clinics damage index; SLEDAI-2K = systemic lupus erythematosus disease activity index 2000; ACR= American College of Rheumatology.

| | **NPSLE (79)** | **non-NPSLE (142)** | **NPSLE vs non-NPSLE p-value** |
|---|---|---|---|
| **sex** female | 74 (93%) | 133 (93%) | 0.77 |
| **age** | 40.0 (24-49) | 40.5 (24-57) | 0.24 |
| **disease duration (years)** | 7.0 (0.2-20.2) | 6.8 (0.4-18.0) | 0.56 |
| **age of onset** | 27 (16-42) | 31 (16-49) | 0.07 |
| **SDI score** | 1 (0-2) | 0 (0-2) | 0.10 |
| **SLEDAI2k score** | 3 (0-14) | 3 (0-8) | 0.37 |
| **ACR criteria** | 3 (4-7) | 5 (4-7) | 0.68 |
| **WMH volume** in ml | 0.105 (0-1.67) | 0.076 (0-1.81) | 0.46 |
| **WMH number** | 2 (0-11.0) | 1 (0-10.9) | 0.13 |

**Supplementary Table S4. Demographic and clinical data for each cluster.** Data are represented as number (percentage) or median (10-90 percentile). Differences between the five different clusters and the SLE without detectable WMH are expressed in p-value and calculated for nominal variables with Chi-square tests (sex) and for continuous variables, based on their not-normally distribution, with Kruskal-Wallis test. Pairwise comparison was calculated using the Mann-Whitney-U test, whereas post-hoc analysis using Bonferroni correction showed significances between the following groups: age: 5 ≠ no WMH; WMH volume in ml: 3 ≠ 4; WMH number: 2 ≠ 3; 3 ≠ 4.
* p<0.05
SDI= systemic lupus international collaborating clinics damage index; SLEDAI-2K = systemic lupus erythematosus disease activity index 2000; ACR= American College of Rheumatology.

| | **1** | **2** | **3** | **4** | **5** | **no WMH** | **p-value** |
|---|---|---|---|---|---|---|---|
| **sex** female | 48 (92.3%) | 53 (93.0%) | 22 (95.7%) | 28 (93.3%) | 21 (87.5%) | 35 (100.0%) | 0.511 |
| **age** | 36.0 (24.0-49.7) | 44.0 (24.6-60.4) | 38.0 (28.6-50.0) | 41.5 (28.7-52.4) | 45.5 (23.1-58.0) | 35.0 (24.0-44.6) | **0.005*** |
| **disease duration (years)** | 7.0 (0.7-17.9) | 6.6 (0.4-23.4) | 8.2 (0.2-17.0) | 9.0 (0.4-23.0) | 6.75 (0.3-15.1) | 6.3 (0.8-19.1) | 0.949 |
| **age of onset** | 26.95 (15.2-42.8) | 30.6 (16.9-55.6) | 29.0 (19.5-43.8) | 30.75 (15.9-45.2) | 39.95 (12.9-53.8) | 24.2 (17.5-41.3) | **0.045*** |
| **SDI score** | 0.0 (0.0-2.0) | 0.0 (0.0-3.0) | 1.0 (0.0-2.0) | 1.0 (0.0-2.1) | 1.0 (0.0-2.0) | 0.0 (0.0-2.0) | 0.353 |
| **SLEDAI2k score** | 2.5 (0.0-9.8) | 2.0 (0.0-8.8) | 2.0 (0.0-7.6) | 4.0 (0.0-8.4) | 2.5 (0.0-12.0) | 4.0 (0.0-13.2) | 0.518 |
| **ACR criteria** | 5.0 (4.0-7.9) | 5.0 (4.0-7.0) | 5.0 (4.0-7.0) | 5.0 (3.9-7.1) | 4.5 (4.0-5.7) | 5.0 (4.0-7.0) | 0.265 |
| **WMH volume** in ml | 0.16 (0.0-0.7) | 0.25 (0.0-4.1) | 0.04 (0.0-0.4) | 0.32 (0.0-1.9) | 0.12 (0.0-7.3) | NA | **0.008*** |
| **WMH number** | 2.0 (0.0-7.9) | 3.0 (0.0-14.4) | 1.0 (0.0-2.8) | 4.0 (1.0-9.4) | 2.0 (0.0-23.0) | NA | **0.003*** |

**Supplementary Table S5. White matter hyperintensity volumes in each white matter tract across the clusters.** White matter hyperintensity (WMH) volumes (in ml) are expressed as median (10-90 percentile). Each row represents one white matter tract. Each column represents one cluster. Grey cells represent the tract where there is the highest WMH volume in each cluster. Differences between the clusters are expressed in p-value and calculated, based on their not-normally distribution, with Kruskal-Wallis tests.
R=right; L=left.
*= p value<0.05

| | **Cluster 1** | **Cluster 2** | **Cluster 3** | **Cluster 4** | **Cluster 5** | **p-value** |
|---|---|---|---|---|---|---|
| **Anterior thalamic radiation L** | 0.29 (0.0-13.8) | 3.35 (0.0-97.4) | 0.0 (0.0-1.8) | 22.52 (4.3-87.3) | 1.3 (0.0-212.6) | **0.000*** |
| **Anterior thalamic radiation R** | 0.05 (0.0-14.0) | 27.55 (3.1-343.4) | 0.0 (0.0-3.1) | 8.91 (0.0-51.8) | 0.78 (0.0-154.9) | **0.000*** |
| **Corticospinal tract L** | 0.0 (0.0-0.1) | 0.0 (0.0-0.8) | 0.0 (0.0-0.0) | 0.0 (0.0-0.1) | 0.0 (0.0-4.6) | **0.013*** |
| **Corticospinal tract R** | 0.0 (0.0-0.1) | 0.0 (0.0-0.9) | 0.0 (0.0-0.0) | 0.0 (0.0-0.0) | 0.0 (0.0-0.9) | 0.325 |
| **Cingulum cingulate gyrus L** | 0.0 (0.0-0.0) | 0.0 (0.0-0.0) | 0.0 (0.0-0.0) | 0.0 (0.0-0.0) | 0.0 (0.0-0.4) | 0.206 |
| **Cingulum cingulate gyrus R** | 0.0 (0.0-0.0) | 0.0 (0.0-0.2) | 0.0 (0.0-1.7) | 0.0 (0.0-0.0) | 0.0 (0.0-0.1) | **0.023*** |
| **Cingulum hippocampus L** | 0.0 (0.0-0.9) | 0.0 (0.0-0.5) | 0.0 (0.0-0.0) | 0.0 (0.0-0.1) | 0.0 (0.0-0.1) | **0.010*** |
| **Cingulum hippocampus R** | 0.0 (0.0-0.2) | 0.0 (0.0-0.0) | 0.0 (0.0-0.0) | 0.0 (0.0-0.0) | 0.0 (0.0-1.8) | **0.043*** |
| **Forceps major** | 24.56(1.7-123.8) | 3.39 (0.0-110.1) | 0.0 (0.0-3.2) | 0.3 (0.0-13.2) | 4.8 (0.0-86.0) | **0.000*** |
| **Forceps minor** | 1.4 (0.0-13.3) | 10.21 (0.1-66.6) | 7.43 (0.9-25.4) | 14.29 (0.4-42.7) | 1.48 (0.0-63.4) | **0.000*** |
| **Inferior fronto-occipital fasciculus L** | 2.54 (0.0-16.9) | 0.86 (0.0-46.5) | 0.0 (0.0-1.2) | 4.3 (0.0-23.7) | 1.12 (0.0-146.1) | **0.000*** |
| **Inferior fronto-occipital fasciculus R** | 2.12 (0.0-23.8) | 7.63 (0.3-76.3) | 0.0 (0.0-0.5) | 2.22 (0.0-19.3) | 9.15 (0.0-210.9) | **0.000*** |
| **Inferior longitudinal fasciculus L** | 0.97 (0.0-5.4) | 0.0 (0.0-16.8) | 0.0 (0.0-0.0) | 0.0 (0.0-2.1) | 0.32 (0.0-55.2) | **0.000*** |
| **Inferior longitudinal fasciculus R** | 1.44 (0.0-7.2) | 0.0 (0.0-14.7) | 0.0 (0.0-0.0) | 0.0 (0.0-0.8) | 1.36 (0.0-30.7) | **0.000*** |
| **Superior longitudinal fasciculus L** | 0.0 (0.0-0.3) | 0.0 (0.0-16.3) | 0.0 (0.0-0.0) | 0.0 (0.0-0.7) | 1.8 (0.0-173.1) | **0.000*** |
| **Superior longitudinal fasciculus R** | 0.0 (0.0-0.2) | 0.0 (0.0-14.3) | 0.0 (0.0-0.0) | 0.0 (0.0-1.3) | 0.16 (0.0-143.9) | **0.000*** |
| **Uncinate fasciculus L** | 0.03 (0.0-3.5) | 0.63 (0.0-14.5) | 0.18 (0.0-0.6) | 2.88 (0.0-15.9) | 0.34 (0.0-44.8) | **0.000*** |
| **Uncinate fasciculus R** | 0.0 (0.0-2.0) | 1.77 (0.0-17.0) | 0.0 (0.0-0.4) | 0.62 (0.0-6.4) | 0.09 (0.0-20.3) | **0.000*** |
| **Superior longitudinal fasciculus temporal L** | 0.0 (0.0-0.1) | 0.0 (0.0-5.7) | 0.0 (0.0-0.0) | 0.02 (0.0-0.4) | 0.3 (0.0-70.9) | **0.000*** |
| **Superior longitudinal fasciculus temporal R** | 0.0 (0.0-0.0) | 0.0 (0.0-3.5) | 0.0 (0.0-0.0) | 0.0 (0.0-0.2) | 0.0 (0.0-68.8) | **0.000*** |

**Supplementary Table S6. Distribution of white matter hyperintensity volumes in different white matter tracts across SLE subgroups.** White matter hyperintensity (WMH) volumes (in ml) in NPSLE and non-NPSLE patients are expressed as median (10-90 percentile). Each row represents one white matter tract. Differences between patient's groups are expressed in p-value and calculated with Mann-Whitney U tests, based on their not-normally distribution.
NPSLE= neuropsychiatric systemic lupus erythematosus; R= right; L= left.
*= p value<0.05

| | **NPSLE** | **non-NPSLE** | **p-value** |
|---|---|---|---|
| **Anterior thalamic radiation L** | 1.43 (0.0-52.2) | 0.16 (0.0-70.6) | 0.156 |
| **Anterior thalamic radiation R** | 3.52 (0.0-111.1) | 0.08 (0.0-97.6) | **0.024*** |
| **Corticospinal tract L** | 0.0 (0.0-0.3) | 0.0 (0.0-0.2) | 0.828 |
| **Corticospinal tract R** | 0.0 (0.0-0.3) | 0.0 (0.0-0.0) | 0.838 |
| **Cingulum cingulate gyrus L** | 0.0 (0.0-0.0) | 0.0 (0.0-0.0) | 0.087 |
| **Cingulum cingulate gyrus R** | 0.0 (0.0-0.0) | 0.0 (0.0-0.0) | 0.597 |
| **Cingulum hippocampus L** | 0.0 (0.0-0.3) | 0.0 (0.0-0.5) | 0.530 |
| **Cingulum hippocampus R** | 0.0 (0.0-0.1) | 0.0 (0.0-0.0) | 0.072 |
| **Forceps major** | 3.39 (0.0-76.4) | 0.4 (0.0-67.7) | 0.050 |
| **Forceps minor** | 4.06 (0.0-43.0) | 1.57 (0.0-39.6) | 0.131 |
| **Inferior fronto-occipital fasciculus L** | 1.27 (0.0-15.7) | 0.06 (0.0-27.6) | 0.132 |
| **Inferior fronto-occipital fasciculus R** | 2.24 (0.0-29.3) | 0.4 (0.0-28.4) | **0.010*** |
| **Inferior longitudinal fasciculus L** | 0.0 (0.0-5.1) | 0.0 (0.0-13.3) | 0.364 |
| **Inferior longitudinal fasciculus R** | 0.11 (0.0-6.6) | 0.0 (0.0-7.2) | **0.041*** |
| **Superior longitudinal fasciculus L** | 0.0 (0.0-1.4) | 0.0 (0.0-3.4) | 0.364 |
| **Superior longitudinal fasciculus R** | 0.0 (0.0-1.4) | 0.0 (0.0-5.7) | 0.481 |
| **Uncinate fasciculus L** | 0.3 (0.0-10.0) | 0.03 (0.0-13.4) | 0.232 |
| **Uncinate fasciculus R** | 0.09 (0.0-12.4) | 0.0 (0.0-11.2) | **0.033*** |
| **Superior longitudinal fasciculus temporal part L** | 0.0 (0.0-0.6) | 0.0 (0.0-1.4) | 0.477 |
| **Superior longitudinal fasciculus temporal part R** | 0.0 (0.0-0.2) | 0.0 (0.0-0.5) | 0.617 |
